# Smoking does not alter treatment effect of intravenous thrombolysis in mild to moderate acute ischemic stroke – a Dutch String-of-Pearls Institute (PSI) Stoke Study

**DOI:** 10.1101/2020.04.17.20056622

**Authors:** Anna Kufner, Martin Ebinger, Gert Jan Luijckx, Matthias Endres, Bob Siegerink, Dutch String-of-Pearls Stroke Study Group

## Abstract

**Background:** The smoking-thrombolysis paradox refers to a better outcome in smokers who suffer from acute ischemic stroke (AIS) following treatment with thrombolysis. However, studies on this subject have yielded contradictory results and an interaction analysis of exposure to smoking and thrombolysis in a large, multicenter database is lacking.

**Methods:** Consecutive AIS patients admitted within 12 hours of symptom onset between 2009 and 2014 from the prospective, multicenter stroke registry (Dutch String-of-Pearls Stroke Study) were included for this analysis. We performed a generalized linear model for functional outcome three months post-stroke depending on risk of the exposure variables (smoking yes/no, thrombolysis yes/no). The following confounders were adjusted for: smoking, hypertension, atrial fibrillation, stroke severity, and stroke etiology.

**Results:** Out of 468 patients, 30.6% were smokers and median baseline NIHSS was 3 (interquartile range 1-6). Smoking alone had a crude and adjusted relative risk (RR) of 0.99 (95% CI 0.89-1.10) and 0.97 (95%CI 0.87-1.01) for good outcome (modified Rankin Score ≤2), respectively. A combination of exposure variables (smoking and thrombolysis) did not change the results significantly (crude RR 0.87 [95% CI 0.74-1.03], adjusted RR 1.1 [95%CI 0.93-1.33]). Smoking alone had an adjusted RR of 1.2 (95% CI 0.6-2.7) for recanalization following thrombolysis (N=88).

**Conclusions:** In patients with mild to moderate AIS admitted within 12 hours of symptom onset, smoking did not modify treatment effect of thrombolysis.

## Introduction

The so-called smoking-paradox of an improved outcome following thrombolysis was first described in smokers with myocardial infarction^1,2^. This phenomenon has resurfaced as a topic of interest in the scientific community as a handful of recent studies have reported similar observations in acute ischemic stroke patients treated with recombinant tissue plasminogen activator (r-tPA) as well as endovascular therapies.^3–6^

The mechanism underlying the pathophysiology of the smoking-thrombolysis paradox remains unclear. Some attribute the observed phenomenon to a systematic lack of adjustment of confounding factors i.e. lower clinical risk profiles of smokers due to younger age and fewer comorbidities.^7,8^ Others argue that there is substantial evidence supporting an alteration of clot dynamics^9,10^ caused by smoke exposure leading to enhanced tPA efficacy in patients with this risk factor.

As of yet, the studies on this subject have yielded contradictory results^3–8,11,12^. Most likely, there is a cumulative effect of younger age, lower clinical risk profiles and more aggressive treatment effect that account for the smoking-thrombolysis paradox. However, a large comprehensive interaction analysis of exposure to smoking and treatment with thrombolysis is still lacking. Therefore, we set out to determine the measures of interaction of smoking status (current smokers vs. non-smokers) and treatment (thrombolysis vs. no thrombolysis) and their attributable risk in terms of functional recovery three month post-stroke and recanalization rates in a large, national multicenter stroke registry.

## Methods

### Patients

All data comes from the Dutch String-of-Pearls Stroke Study - a prospective, multicenter cohort study in which patients with recent stroke presenting within one-month after symptom onset are eligible for enrollment following informed consent^13^. Local ethics committees of all participating hospitals approved the study and all patients provided written informed consent. Research with this data was performed in accordance with the Medical Research Involving Human Subjects Act and codes developed by the Dutch Federation of Medical Scientific Societies. Consecutive acute ischemic stroke patients between October 2009 and October 2014 were included for this analysis. Major inclusion criteria for the current study include the following: ischemic stroke determined via CT or MRI, known time of symptom onset, admission within 12h of symptom onset, and documented smoking status at the time of the event. Patients were excluded from the analysis if they received endovascular therapies or if endovascular treatment status was unknown or undocumented. A flow diagram shows the patient selection criteria for this sub-study (Figure 1).

**Figure 1:**
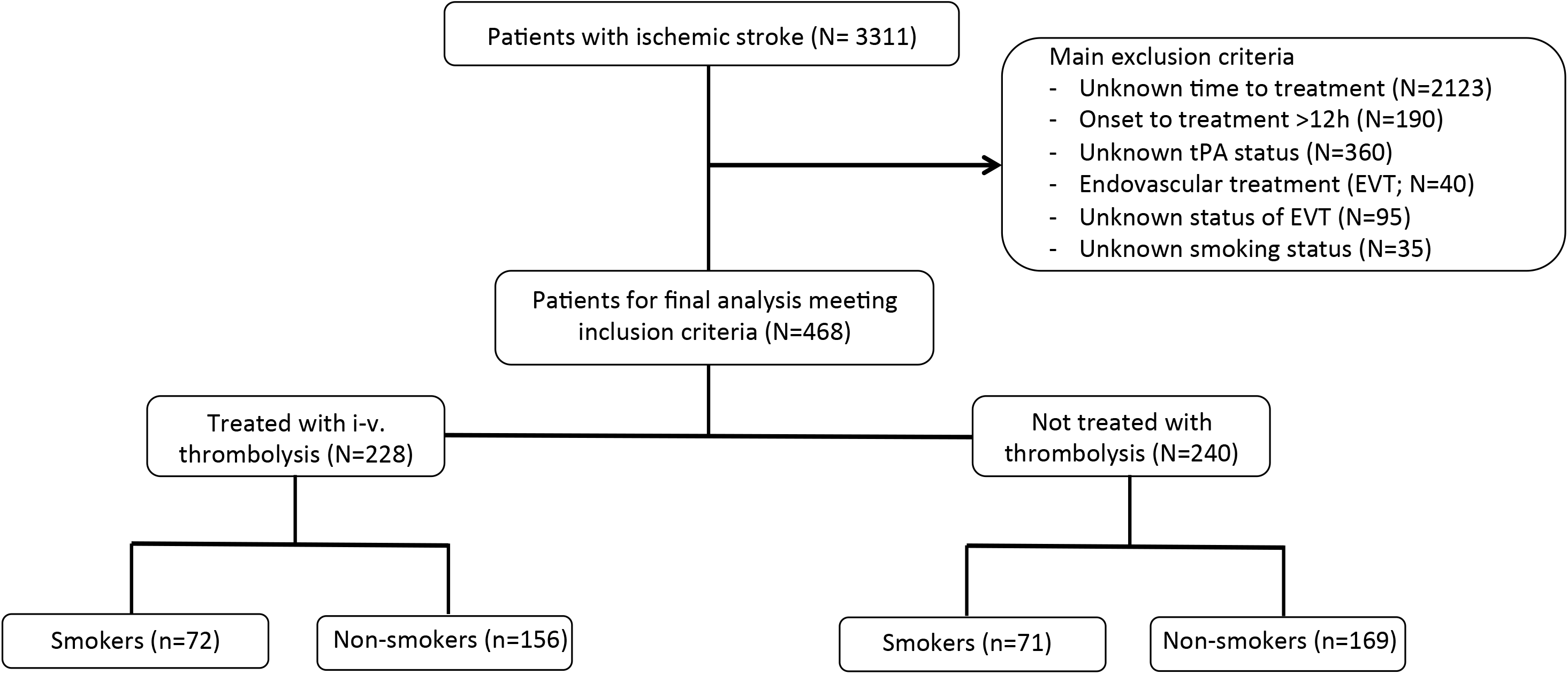
Flow-chart diagram illustrating patient selection for current analyses.

### Regression analyses

We performed several generalized linear regression models with robust estimation of standard errors to estimate relative risks for good functional outcome (mRS ≤2), excellent functional outcome (mRS dichotomized at ≤1), and mortality three months post-stroke depending on risk of the exposure variables (smoking yes/no, thrombolysis yes/no). The two aforementioned exposure variables create four patient groups with combined exposures of smoking and treatment (i.e. -/-, -/+, +/- and +/+). Crude and adjusted relative risk (RRs) were calculated for the four patient groups based on the exposures. Adjusted RRs include correction with the following variables for functional recovery: smoking status, hypertension, atrial fibrillation, stroke severity (NIHSS) and stroke etiology.

## Results

### Patient characteristics

468 patients were included in the final analysis; 30.6% were smokers. For a detailed description of basic demographics and baseline parameters of the entire study group, as well as sub-groups based on smoking-status, refer to Table 1. Compared to non-smokers, smokers had lower rates of hypertension and atrial fibrillation. Stroke etiology differed significantly between groups; smokers presented with higher rates of large-artery atherosclerotic strokes compared to non-smokers who presented with higher rates of cardioembolic stoke. Infarct localization and stroke severity did not differ among groups (Table 1).

**Table 1:** Basic demographics and baseline clinical parameters of the entire cohort, and sub-groups based on smoking status. All variables had <5% missing, with TOAST having the largest percentage of missings (i.e. 4.5%). SD= standard deviation; NIHSS = National Institute of Stroke Scale; IQRL = interquartile range limit; ACA = Anterior cerebral artery; PCA = Posterior cerebral artery; MCA = Middle cerebral artery; TOAST = Trial of Org 10172 in Acute Stroke Treatment.

|  | **All patients**<br>**(N=468)** | **Smokers**<br>**(N=143)** | **Non-smokers**<br>**(N=325)** |
| --- | --- | --- | --- |
| Age, mean (SD) | 66.2 (13.8) | 58.9 (12.7) | 69.3 (13.0) |
| sex, % female | 44.0 (206) | 44.1 (63) | 44 (143) |
| Cerebrovascular risk factors |  |  |  |
| Diabetes mellitus, % (n) | 16.8 (78) | 12.1 (17) | 18.8 (61) |
| Hyperlipidemia, % (n) | 26.1 (119) | 24.1 (34) | 27.0 (85) |
| Atrial fibrillation, %(n) | 12.1 (56) | 6.4 (9) | 14.6 (47) |
| Hypertension, % (n) | 50.1 (231) | 40.9 (58) | 54.2 (173) |
| Baseline NIHSS, median (IQRL) | 3 (1-6) | 3 (1-7) | 2 (1-6) |
| Time to treatment, median (IQRL) | 105 (75 – 150) | 100 (72-143) | 110 (75-155) |
| Thrombolysis, %(n) | 48.7 (228) | 50.3 (72) | 48.0 (156) |
| Infarction in arterial territories |  |  |  |
| ACA, %(n) | 39.1(173) | 41.0(55) | 38.2 (118) |
| PCA, %(n) | 7.3 (32) | 4.5 (6) | 8.4 (26) |
| MCA, %(n) | 23.5 (105) | 26.5 (36) | 22.2(69) |
| Basilar artery, %(n) | 9.0 (40) | 9.8 (13) | 8.7 (27) |
| Vertebral artery, %(n) | 4.5 (20) | 1.5(2) | 5.8(18) |
| Stroke etiology (TOAST) |  |  |  |
| Large-artery atherosclerosis, %(n) | 23.5 (105) | 24.8(33) | 22.9 (72) |
| Cardioembolism, %(n) | 23.5 (88) | 11.3 (15) | 23.3 (73) |
| Small vessel occlusion, %(n) | 19.7 (85) | 27.1 (36) | 15.6 (49) |
| Other, %(n) | 6.7 (30) | 7.5 (10) | 6.4 (20) |
| Undetermined, %(n) | 31.1 (139) | 29.3 (39) | 31.9 (100) |

### Generalized linear model for functional recovery and recanalization

Comparing smokers and non-smokers revealed no significant differences in terms of good outcome of mRS≤2 (70.6% vs. 72.9%), excellent outcome of mRS≤1 (46.2% vs. 55.4%), or mortality (4.2% vs. 4.9%). Smoking status had a crude and adjusted RR of 0.99 (95% CI 0.89-1.11) and 0.97 (95% CI 0.87-1.08) for a good outcome, respectively. In a sub-group analysis including only patients with documented recanalization status (N=88), smoking had a crude RR of 1.3 (95% CI 0.6-2.7 and an adjusted RR (including age, time-to-treatment, stroke etiology) of 1.2 (95% CI 0.6-2.7) for recanalization. Results from the combined exposure analyses (i.e. Smoking + thrombolysis) for a good outcome (mRS≤2), excellent outcome (mRS≤1), and mortality three months following the index event are presented in Table 2. In short, no clear interaction could be observed.

**Table 2:** Relative risks (RR) for a good outcome (modified Rankin Score [mRS]≤2), excellent outcome (mRS≤1), and mortality three months post-stroke, presenting crude and adjusted values (adjusted for presence of hypertension, atrial fibrillation, National Institute of Stroke Scale on admission, stroke etiology categories). Adjustment was not performed for mortality analyses due to low numbers.

| Thrombolysis | Smoking | Number of endpoints/total | Crude RR (95% CI) | Adjusted RR (95% CI) |
| --- | --- | --- | --- | --- |
| *Good outcome mRS≤2* |  |  |  |  |
| - | - | 136/162 | 1 (reference) | 1 (reference) |
| - | + | 55/68 | 0.96 (0.84 -1.10) | 0.92 (0.82 -1.04) |
| + | - | 101/143 | 0.84 (0.74 – 0.95) | 1.06 (0.94 – 1.20) |
| + | + | 46/63 | 0.87 (0.74 – 01.03) | 1.11 (0.93-1.33) |
| *Excellent outcome mRS≤1* |  |  |  |  |
| - | - | 105/162 | 1 (reference) | 1 (reference) |
| - | + | 39/68 | 0.88 (0.70 -1.12) | 0.88 (0.69 – 1.10) |
| + | - | 75/143 | 0.81 (0.67 – 0.98) | 1.11 (0.90 – 1.37) |
| + | + | 27/63 | 0.66 (0.49 – 0.90) | 0.95 (0.68 – 1.34) |
| *Mortality* |  |  |  |  |
| - | - | 8/162 | 1 (reference) | - |
| - | + | 4/68 | 1.29 (0.4 – 3.8) | - |
| + | - | 8/143 | 1.1 (0.4 – 2.9) | - |
| + | + | 2/63 | 0.6(0.1-3.0) | - |

## Discussion

We observed no biological interaction of smoking and intravenous thrombolysis in terms of functional recovery three months post-stroke in this multicenter cohort of ischemic stroke patients admitted within 12 hours of symptom onset.

Interestingly, both crude and adjusted analyses suggest that smoking alone has no effect on long-term functional recovery post-stroke despite lower clinical risk profiles of these patients, although the precision of these analyses is limited. Yet in our combined exposure analyses, smoking alone had a crude and adjusted RR of 0.88 for an excellent outcome, which is indicative of its harmful effects on long-term outcome alone. The seemingly adverse effect of tPA-administration alone on functional recovery disappeared when confounding factors such as stroke severity were considered (crude RR of 0.84 and adjusted RR of 1.06 compared to crude RR of 0.81 and adjusted RR 1.11 for good and excellent outcome, respectively), which can be explained by confounding by indication. A combination of tPA-administration and smoking did not lead to an improved functional outcome (Table 2). These results are an indicator that smoking status negatively influences outcome, regardless of treatment with thrombolysis in this cohort.

Similar to previous reports^3–5,8^, approximately 31% of patients were smokers. As expected, patients with this risk factor had fewer cerebrovascular comorbidities (hypertension^3,4,11^ and AF^11^), and more frequently suffered from large-artery atherosclerotic strokes^4,11^ (Table 1). Smoking led to increased recanalization rates in patients treated with thrombolysis in this cohort (adjusted RR 1.2 95% CI 0.6-2.7). However, due to small numbers an additive interaction analysis for smoking and thrombolysis for recanalization could not be performed and results should be interpreted with caution.

This cohort included predominately minor strokes (median baseline NIHSS: 3 IQR 1-6). Assuming smoking may modify treatment effect of thrombolysis by increasing recanalization rates of large vessel occlusions, any biological interaction of smoking and tPA may have been missed in this analysis. Previous studies that found an enhanced treatment efficacy in smokers included patients with more severe strokes and higher rates of proven vessel occlusion^3–6^. Further limitations of this study include missing data and lack of information on proven vessel occlusion and exclusion of patients with undocumented time of symptom onset, which may have led to selection bias.

In conclusion, smoking did not modify treatment effect of thrombolysis in acute ischemic stroke patients in this cohort. However, a large, multicenter cohort analysis including patients with proven vessel occlusion and more severe strokes is warranted to further investigate a potential biological interaction between smoking and intravenous and intra-arterial thrombolysis.

## Data Availability

The datasets generated during and/or analysed during the current study are available from the corresponding author on reasonable request.

## References

1. Grines CL, Topol EJ, O’Neill WW, et al. Effect of cigarette smoking on outcome after thrombolytic therapy for myocardial infarction. Circulation. 1995;91(2):298–303. doi:10.1161/01.CIR.91.2.298

2. Barbash GI, White HD, Modan M, et al. Significance of smoking in patients receiving thrombolytic therapy for acute myocardial infarction. Experience gleaned from the International Tissue Plasminogen Activator/Streptokinase Mortality Trial. Circulation. 1993;87(1):53–58. doi:10.1161/01.CIR.87.1.53

3. Kufner A, Nolte CH, Galinovic I, et al. Smoking-thrombolysis paradox: Recanalization and reperfusion rates after intravenous tissue plasminogen activator in smokers with ischemic stroke. Stroke. 2013;44:407–413. doi:10.1161/STROKEAHA.112.662148

4. Kvistad CE, Oeygarden H, Logallo N, Thomassen L, Waje-Andreassen U, Naess H. Is smoking associated with favourable outcome in tPA-treated stroke patients? Acta Neurol Scand. 2014;130(5):299–304. doi:10.1111/ane.12225

5. Meseguer E, Labreuche J, Gonzalez-Valcarcel J, et al. The smoking paradox: impact of smoking on recanalization in the setting of intra-arterial thrombolysis.Cerebrovasc Dis Extra. 2014;4(2):84–91. doi:10.1159/000357218

6. von Martial R, Gralla J, Mordasini P, et al. Impact of smoking on stroke outcome after endovascular treatment. PLoS One. 2018. doi:10.1371/journal.pone.0194652

7. Aune E, Roislien J, Mathisen M, Thelle DS, Otterstad JE. The “smoker’s paradox” in patients with acute coronary syndrome: a systematic review. BMC Med. 2011;9:97. doi:10.1186/1741-7015-9-97

8. Ali SF, Smith EE, Reeves MJ, et al. Smoking Paradox in Patients Hospitalized With Coronary Artery Disease or Acute Ischemic Stroke. Circ Cardiovasc Qual Outcomes. 2015;8(6 suppl 3):S73–S80. doi:10.1161/CIRCOUTCOMES.114.001244

9. Barua RS, Sy F, Srikanth S, et al. Effects of cigarette smoke exposure on clot dynamics and fibrin structure: an ex vivo investigation. Arter Thromb Vasc Biol. 2010;30:75–79. doi:10.1161/ATVBAHA.109.195024

10. Meade TW, Imeson J, Stirling Y. EFFECTS OF CHANGES IN SMOKING AND OTHER CHARACTERISTICS ON CLOTTING FACTORS AND THE RISK OF ISCHAEMIC HEART DISEASE. Lancet. 1987;330(8566):986–988. doi:10.1016/S0140-6736(87)92556-6

11. Lee J-H, Lee JY, Ahn SH, et al. Smoking is Not a Good Prognostic Factor following First-Ever Acute Ischemic Stroke. J stroke. 2015;17(2):177–191. doi:10.5853/jos.2015.17.2.177

12. Schlemm L, Kufner A, Boutitie F, et al. Current Smoking Does Not Modify the Treatment Effect of Intravenous Thrombolysis in Acute Ischemic Stroke Patients—A Post-hoc Analysis of the WAKE-UP Trial. Front Neurol. 2019. doi:10.3389/fneur.2019.01239

13. Nederkoorn PJ, van Dijk EJ, Koudstaal PJ, et al. The Dutch String-of-Pearls Stroke Study: Protocol of a large prospective multicenter genetic cohort study. Int J Stroke. 2015;10(1):120–122. doi:10.1111/ijs.12359

